# Feasibility study of a novel mHealth clinical decision support application to enable Community Health Workers to manage hypertension in rural Guatemala

**DOI:** 10.64898/2026.01.21.26344552

**Authors:** Sean Duffy, Alejandro Chavez, Ian Stanley Arthur, Gabriella Lucia Ortiz, Alvaro Bermudez-Cañete, Pablo Nuñez-Perez, Elizabeth White, Valerie Aguilar, Juan Aguirre, Do Dang, Celina Sonia Perez Abaj, Yoselin Emelina Letona López, Rafael Tun, Taryn McGinn Valley

## Abstract

**Background:** Hypertension is a leading global cause of morbidity and mortality, especially in low- and middle-income countries (LMICs), where low health service access limits diagnosis and treatment. Community health workers (CHWs) supported by mobile health (mHealth) clinical decision support (CDS) tools may help. This study evaluated the feasibility of CHW-led hypertension management using an mHealth CDS application in rural Guatemala.

**Methods:** We conducted a six-month, single-group feasibility study in rural San Lucas Tolimán, Guatemala. Trained CHWs used a CommCare-based CDS application wherein algorithms based on WHO HEARTS guided medication titration, lifestyle counseling, and physician consultation. Adults (≥18 years) with diagnosed hypertension were followed monthly for six months. The primary feasibility outcome was prescribing agreement between CHWs and supervising physicians with application antihypertensive recommendations (minimum acceptable agreement = 90%). Primary clinical outcomes were changes in systolic (SBP) and diastolic (DBP) blood pressure. Secondary outcomes included retention, visit completion, patient satisfaction, and safety.

**Results:** Thirty-two participants were enrolled (84% female); 30 (93.8%) completed six-month follow-up, with 96.4% of possible visits completed. Overall CHW-physician agreement with application recommendations was 96.7%, increasing to 98.9% over the final three months. Median SBP decreased 7.5 mmHg (95% CI 1.0-12.0) and mean DBP 3.1 mmHg (95% CI 0.1-6.1). The proportion of patients with controlled SBP (<140 mmHg) increased from 66.7% to 76.7% (p = 0.505). Eleven application errors (5% of 217 visits) occurred early and were corrected. There were no adverse events requiring hospitalization. Patient satisfaction remained high.

**Conclusions:** In this pilot, CHWs supported by an mHealth CDS application safely and effectively managed hypertension with high physician agreement, patient retention, and blood pressure improvement. These findings demonstrate the feasibility of CHW task-sharing hypertension management in low-resource settings and support evaluation of this approach in larger trials

**Registration:** This study was registered with ClinicalTrials.gov (ID NCT05479097).

## Background

Hypertension is the most common chronic disease in adults and is a leading global cause of morbidity and mortality. Around a third of all adults, approximately 1.4 billion people, are estimated to have hypertension globally.[1] The prevalence of hypertension, along with the rate of hypertension-related deaths, is now higher in low- and middle-income countries (LMICs) than in high-income countries.[1] Many patients are able to effectively treat and manage their hypertension through lifestyle changes and affordable medications. However, successful management requires continuous monitoring and interaction with health services.[2] Lack of access to skilled clinicians and medications contributes to a higher prevalence of uncontrolled hypertension and premature mortality in LMICs.[1] Given the magnitude of hypertension as a public health problem, increasing access to effective treatment needs to be a global health priority.[3]

Community health workers (CHWs) are frontline public health workers who can serve as a vital link between communities and health systems, connecting people to information, resources, and services.[4] Previous studies have shown that CHWs can effectively aid hypertension management within health systems in LMICs.[5] CHWs’ involvement in hypertension programs has primarily been in supportive and educational roles, with limited evidence for their direct involvement in care provision.[6] LMICs lack treatment infrastructure and skilled providers to treat hypertension, limiting their capacity to address the rise in chronic non-communicable diseases such as hypertension. Continued investment in integrated care is crucial to overcome this problem and reduce morbidity and premature mortality.[7–9] There is currently insufficient evidence regarding how CHWs can fill this gap in hypertension care, including whether mHealth tools are feasible for CHWs to use with remote supervision, and if these approaches could provide adequate support and management of patients.

With this study, in rural areas of Guatemala with poor primary care infrastructure, we aim to address the gap in research about direct hypertension management through CHWs and mHealth tools. To strengthen this scholarship, we performed a pilot study to show the feasibility of this approach. By proving the feasibility of CHW-led hypertension management enabled by a mobile application, our study facilitates future testing of this approach in a larger trial.

## Methods

### Study setting

San Lucas Tolimán (SLT) is a rural municipality in the western highlands of Guatemala, located on the shores of Lake Atitlan. The town proper has a population of 35,000, with another ∼20,000 living in small surrounding communities. The majority of inhabitants in SLT are Indigenous Kaqchikel Maya. The most common sources of income are agricultural labor from coffee or banana plantations or informal labor such as weaving. The study was conducted in the rural communities of SLT with monthly visits occurring in community spaces, churches, or classrooms. The leading Guatemalan organization for this project was the San Lucas Mission (SLM), a non-profit organization providing healthcare and other social services to the people of SLT and neighboring municipalities. SLM sponsors a CHW network serving the rural communities of SLT.

### Mobile application development

Over a 12-month period, we adapted our successful clinical decision support (CDS) application, designed to assist CHWs with diabetes management in rural Guatemala[10] to provide guidance for hypertension management and cardiovascular risk reduction. As with our diabetes application, this application was developed on the CommCare platform, which is widely used by frontline health workers in LMICs. Algorithms for titrating antihypertensives embedded in this application (Appendix S1) were based on the WHO HEARTS protocol for hypertension management.[11]

We chose amlodipine, a calcium channel blocker (CCB), enalapril, an angiotensin converting enzyme inhibitor (ACEI), and losartan, an angiotensin II receptor blocker (ARB), for our titration algorithms based on international guidelines.[11,12] Due to lower cost and greater availability in Guatemala and in LMICs,[1,13] we designated an ACEI (enalapril) as the first-line renin-angiotensin-aldosterone system (RAAS) inhibitor, with losartan (an ARB) used for patients with enalapril or other ACEI intolerance or who were taking an ARB at enrollment. The algorithms are capable of initiating and adjusting dosages for amlodipine or enalapril/losartan, used as monotherapy or combination. The application also provides guidance on statin and aspirin treatment for patients with prior history of clinical atherosclerotic cardiovascular disease (ASCVD) and statin treatment for primary prevention in high risk patients according to WHO and consensus US guidelines.[11,14] The statin used for this study was atorvastatin. In addition to these medications, the supervising physician was able to manually add additional antihypertensives, such as hydrochlorothiazide. While the application does not provide recommendations for titration of these medications, it is able to recommend continuing these medications if the patient is not hypotensive and does not report any side effects. The application provides a rationale for medication recommendations so the user can independently verify the decision logic.

In addition to medication titration algorithms, the application also provides guidance for lifestyle counseling, the identification of possible complications of hypertension – including hypertensive emergency, coronary artery disease, heart failure, and stroke – and remote consultation with a supervising physician as needed. Prior to the study, we conducted extensive quality assurance testing with the application. To do so, we first fed dummy data into the application covering all possible scenarios to ensure that all recommendations provided by the app were accurate. Then, we observed CHWs using the application in simulated patient encounters to assess usability, fine-tune clarity of prompts and recommendations, and uncover unanticipated workflow problems and application errors.

### CHW training

To be eligible to participate in the study, CHWs had to have already completed the 24-month basic CHW training program focused on care of common conditions and community health promotion. Eligible CHWs then completed a hypertension-specific curriculum consisting of five six-hour training sessions over a six-month period, focusing on antihypertensive treatment, identification of potential complications or complex scenarios needing physician consultation, lifestyle guidance, and application use. Readiness to participate in the study was assessed by CHWs scoring 80% or higher on a written hypertension knowledge test and satisfactory performance on a practical examination, during which a trained observer used a standardized rubric to grade the learner’s use of the mobile application to assess and treat a standardized patient.

### Study design, sample, recruitment, and reporting

We conducted a single-group feasibility study. Target enrollment was 30 patients, which we found sufficient to determine the initial feasibility of our diabetes intervention,[15] and falls within the range of recommended sample sizes for feasibility studies.[16] Inclusion criteria were: subjects age 18 and older with physician-confirmed diagnosis of hypertension AND blood pressure (BP) >=140/90 OR currently taking antihypertensives. Exclusion criteria included pregnancy and comorbid conditions leading to life expectancy <1 year. Patients were recruited from the rural communities of SLT, targeting individuals identified as having hypertension in our earlier study of hypertension screening.[17]

We used the CONSORT 2010 extension for pilot and feasibility trials checklist[18] to report study conduct and outcomes (Appendix S2). This study was registered with ClinicalTrials.gov (ID NCT05479097).

### Study procedures

CHWs met with patients monthly, using the CDS application to guide each visit. At each visit, CHWs measured BP and weight, assessed medication adherence and tolerance, evaluated for possible complications of hypertension, provided lifestyle guidance, and dispensed medication. Teleconsultation with a supervising physician was available as needed during study visits.

Following each visit, application data was uploaded to the secure CommCare server and a supervising physician reviewed the current care plans, adjusting as needed in coordination with CHWs. Total duration of study participation was six months.

At the enrollment and six-month follow-up visits, a fasting blood draw was completed by venipuncture to measure serum creatinine, potassium, lipid profile, glucose, and alanine aminotransferase (ALT). At these visits, patients also completed validated Spanish versions of the Hypertension Self-Care Activity Level Effects (H-SCALE)[19], the Patient Assessment of Care for Chronic Conditions (PACIC)[20], and a single item measure of global patient satisfaction.[21]

To assess patient perspectives, a qualitatively trained researcher and a CHW conducted interviews with each patient after the six-month study visit. The team used a semi-structured interview guide (which was collaboratively developed with the lead researcher, qualitatively trained team member, and CHW team) to allow for some standardization while also enabling patients to bring up any issues or suggestions (see Appendix S3 for the interview guide).

### Outcome measures

#### Primary outcomes

The primary feasibility outcome was clinician agreement with the monthly antihypertensive prescription recommended by the CDS application. We assessed this as the proportion of visits for which both the CHW conducting the visit and the physician reviewing the data after the visit agreed with the antihypertensive recommendations provided by the application. We used this as our primary feasibility metric because we felt that this approach would reflect the overall reliability of the CDS provided by the application. A low level of agreement would indicate the need for frequent consultation between CHWs and physicians during visits and medication changes between visits, increasing workload and decreasing the feasibility of the intervention.

We set a target threshold of at least 90% agreement. While there is no widely-agreed-upon standard for this measure in such programs, studies of successful CDS applications used by non-physician health workers (NPHWs) have found analogous agreement proportions of greater than 90%.[8,10] We assessed agreement over the final 3 months of the trial, as we anticipated that agreement would improve throughout the trial with iterative application improvements.

Primary clinical outcomes included change in systolic blood pressure (SBP), change in diastolic blood pressure (DBP), and change in hypertension control status (defined as SBP < 140 mmHg). Unless otherwise noted, all clinical outcomes were assessed at 6 months.

#### Secondary outcomes

Secondary feasibility outcomes included subject retention, visit completion, and patient global satisfaction and PACIC scores. Feasibility assessment also included usability surveys and feedback from the CHWs who participated in the study, which we have described elsewhere.[22]

Secondary clinical outcomes included changes in weight, serum creatinine (and calculated eGFR), lipid profile, glucose, hypertension self-care activities (H-SCALE scores), and medication adherence. When available, pill count adherence ratio (PCAR)[23] was used to determine adherence, with self-reported adherence used only when PCAR was unavailable. Acceptable adherence was defined as PCAR ≥ 75%, or, when using self-report, reporting taking medications as prescribed at least 5 of 7 days/week. As pill count was not available at baseline because patients had not yet been dispensed medication, we used self-report to determine baseline adherence. We compared baseline adherence to adherence at 6 months, as well as comparing adherence between month 1 and month 6 for cases in which PCAR was available at both timepoints.

#### Safety outcomes

Safety outcomes included the incidence of incorrect recommendations provided by the CDS application and other application errors or malfunctions, as well as the incidence of complications of hypertension or antihypertensive treatment, including significant increase in serum creatinine (≥1.5 x baseline), acute coronary syndrome, cerebrovascular accident, decompensated heart failure, hyperkalemia, and syncope.

#### Statistical analysis

As this study focused on determining the feasibility of CHW-led hypertension management, power analyses were not performed.

For continuous outcomes, we used the Shapiro-Wilk test to determine normality. We summarized normally distributed variables using means and standard deviations, and non-normally distributed variables using medians and interquartile range. We assessed changes in paired measures from baseline to 6 months versus a null hypothesis of no change using paired t-tests (for normally distributed variables) or Wilcoxon signed-rank tests (for non-normally distributed variables). For dichotomous outcomes, we calculated raw proportions for baseline and follow-up periods. We assessed association between the outcome and the two periods versus a null hypothesis of no association using McNemar’s test.

## Results

### Participants

A total of 32 subjects participated in the study; baseline demographic and clinical characteristics are summarized in Table 1. Of note, the study population was heavily skewed towards female participants (84%) and a large majority entered already taking antihypertensives (97%); most began with controlled SBP (66%). Recruitment started in February 2023 and ended August 2023.

**Table 1.**
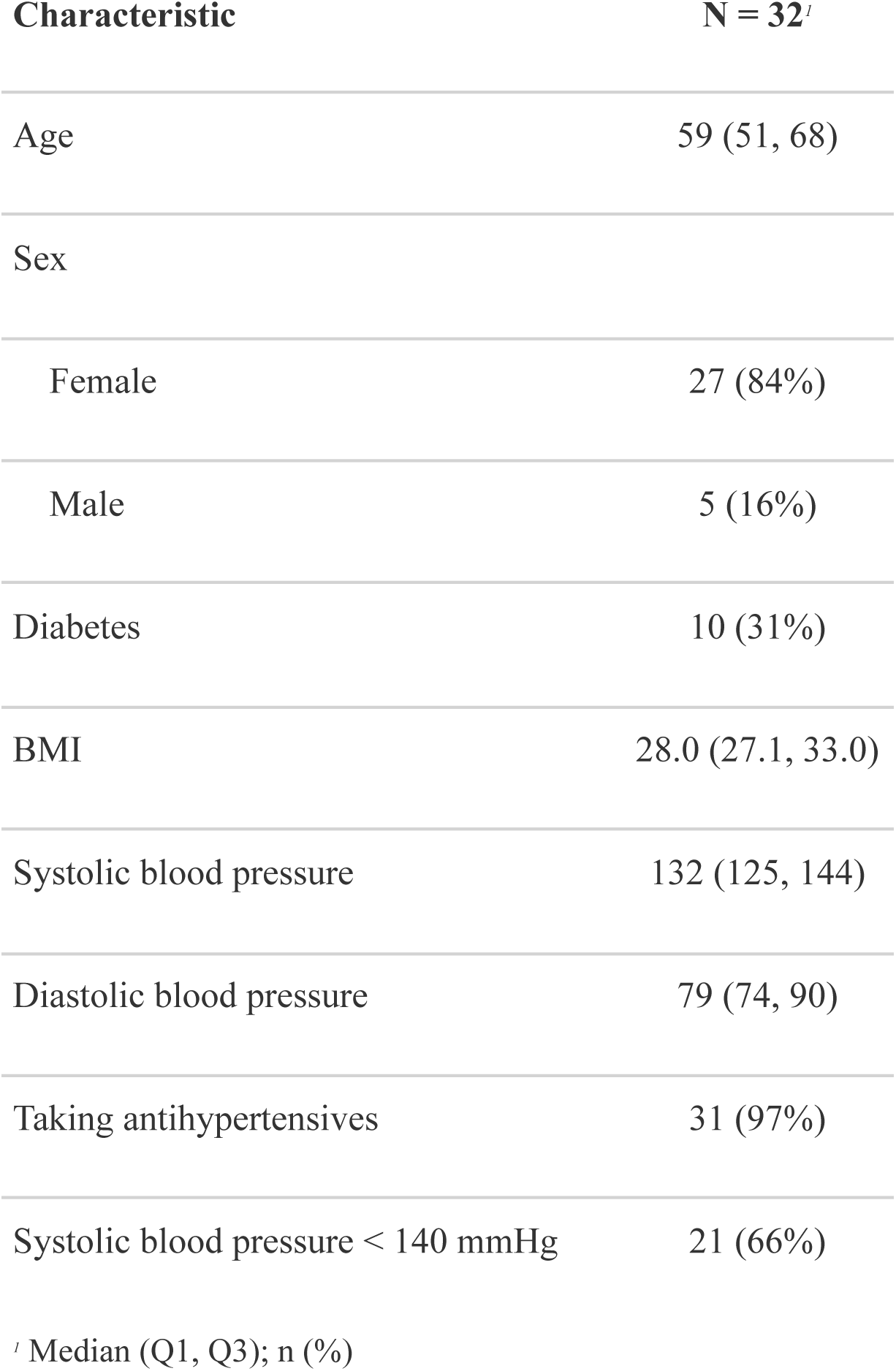
Baseline characteristics of study participants.

| Characteristic | N = 32<sup>1</sup> |
| --- | --- |
| Age | 59 (51, 68) |
| Sex |  |
| Female | 27 (84%) |
| Male | 5 (16%) |
| Diabetes | 10 (31%) |
| BMI | 28.0 (27.1, 33.0) |
| Systolic blood pressure | 132 (125, 144) |
| Diastolic blood pressure | 79 (74, 90) |
| Taking antihypertensives | 31 (97%) |
| Systolic blood pressure < 140 mmHg | 21 (66%) |

<sup>1</sup> Median (Q1, Q3); n (%)

### Primary Outcomes

#### Agreement with application antihypertensive recommendations

CHW and physician agreement with antihypertensive medication recommendations was 98.9% (92 of 93 visits) over the last 3 months of the trial, higher than the prespecified feasibility threshold of 90% agreement. Agreement throughout the whole trial period was 96.7% (210 of 217 visits), also higher than the threshold. Of note, these agreement figures also represent physician agreement alone: there were no instances in which CHWs disagreed with the treatment recommendations provided by the application.

#### Blood pressure

Median decrease in SBP was 7.5 mmHg (n = 30, 95% CI 1.0 to 12.0) from enrollment to 6 months. Mean decrease in DBP for this time period was 3.1 mmHg (n = 30, 95% CI 0.1 to 6.1). Figure 1 shows trends in blood pressure during the trial, illustrating the general downward trend for SBP and DBP.

**Figure 1:**
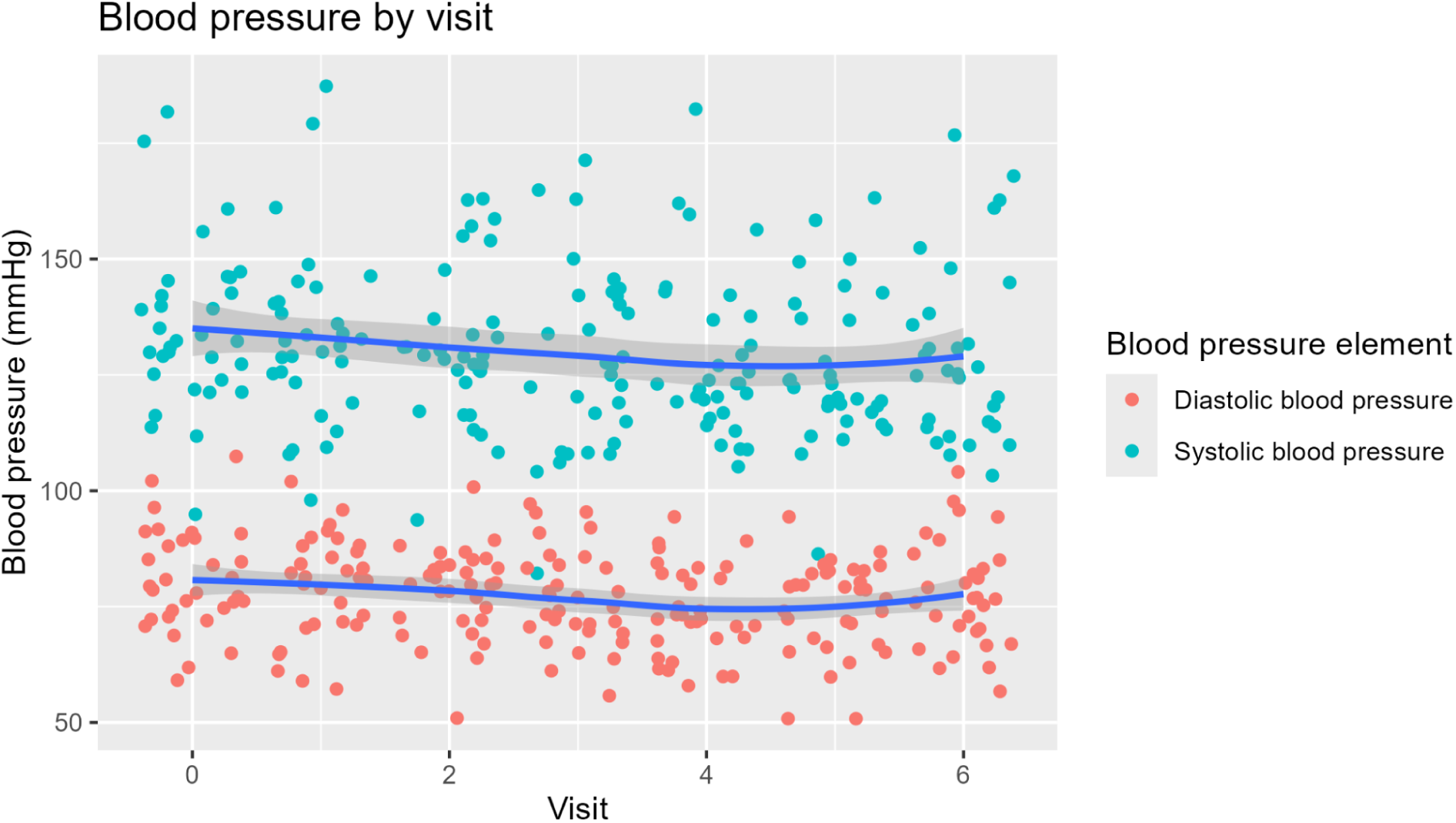
Blood pressure trends during pilot study The percentage of patients with SBP controlled (<140 mmHg) increased from 66.7% at baseline to 76.7% at 6 months (n = 30, McNemar’s chi-squared = 0.444, p = 0.505) (Table 2).

**Table 2:** Blood pressure control status.

| Baseline Control | Control at 6 months |  |  |
| --- | --- | --- | --- |
|  | No | Yes | Total |
| No | 13.3% (4) | 20.0% (6) | 33.3% (10) |
| Yes | 10.0% (3) | 56.7% (17) | 66.7% (20) |
| Total | 23.3 % (7) | 76.7% (23) | 100.0% (30) |

### Secondary Outcomes

#### Subject retention and visit completion

Of the 32 subjects enrolled, 30 (93.8%) remained in the study through the 6 month follow-up visit. One subject withdrew because of relocation to another city and another withdrew due to side effects from antihypertensive medications. Subjects completed 96.4% of possible follow-up visits.

#### Patient satisfaction

Subject ratings on the PACIC survey items were high at baseline, with a median score of 5 (indicating that the desired health care activities were performed “almost always” when they sought hypertension care) for all items. Despite this, there were statistically significant improvements for 6 of the 11 survey items at 6 months, as well as in the average of all included survey items (Table 3).

**Table 3:** PACIC survey results.

|  | Baseline | 6 months |  |
| --- | --- | --- | --- |
| Characteristic | N = 30<sup>1</sup> | N = 30<sup>1</sup> | p-value<sup>2</sup> |
| Asked for my ideas when we made a treatment plan | 5.0 (4.0, 5.0) | 5.0 (4.0, 5.0) | 0.3 |
| Asked to talk about any problems with my medicines or their effects | 5.0 (4.0, 5.0) | 5.0 (5.0, 5.0) | 0.039 |
| Given a written list of things I should do to improve my health | 5.0 (3.0, 5.0) | 5.0 (5.0, 5.0) | 0.007 |
| Satisfied that my care was well organized | 5.0 (5.0, 5.0) | 5.0 (5.0, 5.0) | 0.8 |
| Shown how what I did to take care of my illness influenced my condition | 5.0 (4.0, 5.0) | 5.0 (5.0, 5.0) | 0.025 |
| Helped to set specific goals to improve my eating or exercise | 5.0 (3.0, 5.0) | 5.0 (5.0, 5.0) | 0.005 |
| Given a copy of my treatment plan | 5.0 (3.0, 5.0) | 5.0 (5.0, 5.0) | 0.11 |
| Asked questions, either directly or on a survey, about my health habits | 4.1 (1.2) | 4.6 (0.8) | 0.081 |
| Asked what I would like to discuss about my illness at that visit | 5.0 (3.0, 5.0) | 5.0 (5.0, 5.0) | 0.015 |
| Told how important the things I do to take care of my illness (e.g. exercise) were for my health | 5.0 (4.0, 5.0) | 5.0 (5.0, 5.0) | 0.052 |
| Given a book or monitoring log in which to record the progress I am making | 5.0 (3.0, 5.0) | 5.0 (5.0, 5.0) | 0.029 |
| Average PACIC item score | 4.5 (3.9, 5.0) | 5.0 (4.6, 5.0) | 0.011 |

<sup>1</sup> Median (Q1, Q3); Mean (SD)
<sup>2</sup> Wilcoxon signed rank test with continuity correction; Paired t-test

There was no statistically significant change in global satisfaction with hypertension care from baseline to 6 months, with subjects reporting high satisfaction (median score of 4, “satisfied”, p = 0.71) at both timepoints.

#### Hypertension self-care activities

There were no statistically significant changes in H-SCALE subscores from baseline to 6 months (Table S1). Of note, the alcohol subscore was not included in this analysis because all patients had a score of 0 (no reported alcohol use) at baseline and 6 months.

#### Laboratory and anthropometric outcomes

There was a small but statistically significant increase in creatinine (and associated reduction in eGFR) at 6 months compared to baseline (Table S2). There were also statistically significant reductions in triglycerides, glucose, and ALT. There were no statistically significant changes in BMI or weight from baseline to 6 months (Table S3).

#### Medication adherence

100% of participants were adherent by self-report at baseline, compared to 96% by either adherence measure at six months (n = 28, p = 1). 93% of participants were adherent by PCAR at baseline, which decreased to 79% at six months (n = 14, p = 0.617). (See Tables S4 - S5 for further details)

#### Safety Outcomes

There were 11 application errors resulting in incorrect medication recommendations (5% of 217 total visits). All of these occurred in the first 3 months of the study. One subject experienced a ≥50% increase in creatinine from baseline to 6 months, though creatinine remained within normal limits. Another subject had an episode of syncope related to antihypertensive medications. Regarding complications of hypertension, one subject experienced symptoms (dyspnea on exertion and lower extremity edema) suspected to be related to decompensated heart failure. None of these medication adverse events or hypertension complications required hospitalization.

#### Qualitative Results

Of 32 patients, 25 were at their homes during home visits and interested in participating in an interview. These brief interviews lasted from 3 to 31 minutes, averaging 8 minutes. Interviews were recorded and the qualitative researcher took synchronous notes. She then transcribed several interviews for specific details and listened to all interviews again while analyzing. Common themes were summarized and discussed with the community health workers and research teams.

Overall, patients reported feeling satisfied with and appreciative of the program. In particular, patients appreciated that the care was free, included medicines, and took place close to their homes. Before the pilot program, patients either purchased medication out of pocket or did not take medication. Many took medication only when they could afford it, leading to spotty use, and they often went without.

Summarizing these themes, in a response representative of most interviewees, one woman commented:

> “In my opinion, it [the program] seems good, because it facilitates medicine. When one buys medicine [out-of-pocket], it is expensive, and you have to go into town. And if you don’t have money, there’s no way. Now we’re seeing that we really need medicine, and it helps us have good health. Because for me, this [program] has helped a lot. Before, every month I would buy [medicines], one for [blood] pressure and another for palpitations, and it was getting really expensive. Now, yes, thanks to God, with this support, it has helped me a lot.”

We also asked patients what they would change or improve about the program. A few patients requested treatment for non-hypertension concerns, e.g. allergies and headaches. Several mentioned wanting vitamins. A few, including a midwife, asked for tools to provide medical care, like gloves and wound care supplies.

Overall, though, the majority responded that there was nothing they would change. More than one patient responded that they did not have the “right” to suggest any changes, or, “I don’t know what to say.” In the Guatemalan context, it is quite rare for patients to be asked for feedback about their biomedical care.

## Discussion

In this pilot study of CHW-led hypertension management enabled by an mHealth application providing CDS, we found that this approach is feasible and shows initial indications of clinical safety and efficacy. CHWs were able to make reliable treatment decisions regarding antihypertensive therapy using the application, as the physician reviewing patient data agreed with these decisions greater than 95% of the time, higher than the prespecified feasibility target of 90% agreement. While the primary objective of this study was to demonstrate intervention feasibility, subjects also showed statistically and clinically significant reductions in blood pressure (7.5 mmHg and 3.1 mmHg reductions in SBP and DBP, respectively), despite the fact that all but one subject (97%) entered the study already taking antihypertensives. There were no severe treatment-related adverse events and subjects reported generally high satisfaction with CHW-led care, while we also found improved scores on several measures of the Patient Assessment of Care for Chronic Conditions (PACIC) survey. Taken together, these findings suggest that using mHealth to support task sharing with non-physician health workers for hypertension care is a viable strategy to improve treatment access and outcomes in low-resource settings.

The primary feasibility measure for this study was agreement of CHWs and supervising physicians on the antihypertensive recommendations made by the mHealth application. This metric reflects the ability of CHWs working within this system to function autonomously and effectively at the point of care: high agreement with application recommendations translates to less need to consult with a physician in real time during patient visits, fewer medications changes that need to be coordinated after visits, and improved patient safety due to fewer erroneous or clinically unsound medication prescribing decisions. Agreement throughout the course of this study was 96.7%, higher than the prespecified feasibility threshold of 90%. This rate increased to 98.9% over the final 3 months of the trial, reflecting continuous application improvements based on end-user feedback. Notably, agreement observed in this study is higher than that seen in our earlier study of CHW-led diabetes management (91%)[10] and in the HOPE 4 study (93%), which evaluated a collaborative care model between non-physician health workers and physicians utilizing a CDS mHealth application.[8]

Secondary outcomes were also supportive of intervention feasibility. Subject retention through 6 months was 93.8% and subjects completed 96.4% of possible study visits. This is greater than the median retention of ∼90% reported across a wide variety of clinical trials and commonly targeted retention rates of 85-90%.[24–26] Measures of patient satisfaction were high at baseline. We theorize baseline scores in this medically underserved population may have been inflated by increased access to care in the months prior to the start of this pilot study; all subjects were recruited from our earlier study of hypertension diagnosis[17] and, because the start of the pilot treatment study was delayed, had been provided with prior, bridging antihypertensive medications managed by study physicians. Nevertheless, there were significant improvements in a majority of the PACIC survey items as well as the total PACIC score. The overall rate of application errors leading to incorrect medication recommendations was low, affecting 5% of visits. Though this rate was higher than the <1% error rate observed in our earlier study of a similar diabetes application,[10] all of these errors occurred within the first three months of the study and more than two thirds represented the same error related to statin initiation across multiple patients. This suggests high reliability of the application after early errors were identified and fixed.

The statistically and clinically significant improvements in blood pressure seen at six months in this study compares favorably to those seen in other studies of task sharing interventions for hypertension care in low-resource settings. A 2019 systematic review and meta-analysis of task sharing with non-physician health workers for blood pressure management in LMIC showed mean reductions of 4.85 mmHg SBP and 2.92 mmHg DBP across all interventions, a lower magnitude of improvement than what we observed.[27] BP reductions were more modest for the subset of studies involving CHWs: 3.67 mmHg SBP and 2.29 mmHg DBP.[27]

The HOPE 4 trial employed a very similar model to our intervention, utilizing CHWs equipped with mHealth CDS applications to allow for task sharing with primary care physicians in Colombia and Malaysia.[8] This trial found a greater reduction in mean SBP (20.1 mmHg) and DBP at 6 months in the treatment group (6.1 mmHg) than our intervention. However, baseline BP was significantly higher in the HOPE 4 study cohort (SBP 152.1 mmHg and DBP 84.7 mmHg) than in our study (median SBP 132 mmHg, median DBP 79 mmHg) and therefore, average blood pressure at 6 months was similar for the two studies: 132/78.6 mmHg for HOPE 4 vs 125.5/75.9 mmHg for this study.

Other key differences between HOPE 4 and this study include the autonomy of the CHWs and the complexity of CDS provided by mHealth applications. In HOPE 4, physicians approved all medication changes prior to CHWs carrying them out, a two-step process for medication changes, likely increasing CHW and physician workload and potentially leading to gaps or delays in care. In contrast, supported by the CDS application, CHWs in our study made most medication changes at the point of care using standardized protocols, with retrospective physician review and approval. Additionally, while the HOPE 4 protocols used fixed-dose, single-pill antihypertensive combinations, the advanced decision support provided by the application in our study allowed for initiation and titration of three different antihypertensives, improving personalization of antihypertensive therapy and generalizability to other LMIC settings, where availability and affordability of single-pill combination medications is currently suboptimal.[28,29]

Another trial similar in concept to our study is the China Rural Hypertension Control Project (CRHCP), a cluster RCT randomizing villages to hypertension care delivered by village doctors, a type of CHW operating in rural areas within the Chinese healthcare system, vs usual care.[30] Village doctors were trained on standardized protocols for antihypertensive initiation and titration and were supported and supervised by primary care physicians. Subjects in the intervention group experienced impressive reductions in SBP and DBP at 18 months: 26.3 mmHg and 14.6 mmHg respectively. As in the HOPE 4 study, baseline SBP (157.0 mmHg) and DBP (88.1 mmHg) were significantly higher than in our study, likely accounting for the greater reductions and resulting in similar average BPs at study endpoint in both trials: 130.7/73.5 mmHg in CRHCP vs 125.5/75.9 mmHg for our study. There are also significant differences in the educational background of the village doctors utilized in the CRHCP and the CHWs who carried out the intervention in our study. Nearly 95% of the village doctors had completed at least high school or vocational medical school and more than 60% junior medical college or above. In comparison, only one of the six CHWs who participated in this study had completed any formal education beyond the equivalent of middle school. The use of an mHealth clinical decision application was crucial to the ability of CHWs with less formal education to provide hypertension care, an innovation that was not present in the CRHCP study. We feel that the mHealth strategy increases the potential for wider application of our model across LMIC settings, given that lack of highly educated health workers is one of the key bottlenecks in addressing the rising tide of chronic disease.[31]

There were no statistically significant improvements in hypertension self-care activities, as measured by the H-SCALE questionnaire. Additionally, while baseline medication adherence was exceptionally high (100% by any measure and 93% by PCAR) and did not leave much room for improvement, we saw a non-statistically significant decrease in medication adherence during the course of the study. CHW training in preparation for this study emphasized lifestyle management of hypertension and the mobile application supporting the CHWs provided prompts to conduct patient education on lifestyle changes and medication adherence counseling. However, these findings suggest that more structured and intensive educational interventions may be required to bring about significant improvements in patient hypertension self-care and medication adherence.

We observed a small but statistically significant increase in creatinine (and corresponding reduction in eGFR) during the course of the study: 0.1 mg/dL (10.7 % increase). Rather than representing a true worsening of renal function, we feel that this most likely represents the impact of initiation or escalation of ACEI or ARB therapy during the study, which typically cause a creatinine increase of 10% to 30%, despite long-term renoprotective effects.[32,33] We observed a rate of syncope of 1 event per 16 patient years. In comparison, within the control arm of the SPRINT trial, which had the same SBP goal of <140 mmHg used in this study, the rate of syncopal events was 1 in 206 patient years.[34] Given the small size of our study, we suspect that this difference in rates is primarily due to random chance rather than significantly increased risk of syncope. A larger trial will be necessary to determine the true risk of syncope and other adverse effects of antihypertensive treatment.

Overall, our qualitative data supported that the intervention is both feasible and acceptable to patients. Indeed, they had few negative or constructive comments. However, given the nature of power differentials and healthcare systems in rural Guatemala, it is difficult to conclude that patients actually had no constructive feedback; instead, we suggest that directly asking patients what they would change about programs is likely not the most useful way to elicit program areas for improvement in communities with such explicit power differentials with NGO and foreign healthcare projects. Therefore, while the qualitative data increased our comfort with this pilot intervention, we are revising our qualitative methods for future program evaluation.

An inherent limitation of this study was small sample size, which limits inferences about intervention effectiveness in improving clinical outcomes. Nevertheless, we feel that this sample size was sufficient to demonstrate intervention feasibility and effectiveness is currently being assessed with a much larger, controlled trial.[35] The fact that most participants were receiving regular hypertension care and taking medications at baseline also limited our ability to assess the effects of the intervention on newly-diagnosed patients or those without regular access to care.

However, despite this fact, we did observe clinically significant improvements in blood pressure and other health measures, which supports intervention feasibility.

## Conclusions

In this study of mHealth-enabled hypertension care provided by CHWs, we found that this innovative care model was feasible, safe, and improved blood pressure and other clinical outcomes. We are currently assessing the efficacy of this approach in a cluster randomized, non-inferiority clinical trial comparing direct CHW care with care provided by a physician. The findings from this feasibility study informed refinements that we made to the CHW care model prior to the start of this larger trial, including: the addition of diabetes management functionality to the application used by CHWs, given the high coincidence of diabetes and hypertension in the study cohort; and the inclusion of more structured patient education provided by the CHWs, as well as educational text messaging to patients. We are hopeful that this trial will demonstrate the non-inferiority of this task sharing approach compared to traditional, physician-centric care for hypertension and thus provide another powerful tool for improving patient outcomes in low-resource settings.

## Required statements

### Ethics statement

This research was carried out in accordance with the Declaration of Helsinki. The study was approved by the University of Wisconsin Institutional Review Board (ID # 2022-0794, initial approval 07/02/2022) and by the San Lucas Healthcare Committee (initial approval 09/19/2020), an interdisciplinary, international group which provides local ethics oversight to healthcare projects. Eligible patients provided written informed consent prior to participating in the study.

Study procedures, potential risks and benefits were explained to potential participants by the community health worker (CHW) in Spanish and/or Kaqchikel depending on the patient’s preferred language, and any question answered. A copy of the study information and consent form in Spanish (as patients are generally only literate in Spanish even if they primarily speak Kaqchikel) was provided to the participant. If the CHW obtaining consent was unsure about the patient’s ability to provide informed consent, one of the study supervising physicians was consulted to assess the participant and determine if they could provide consent. This study was registered with ClinicalTrials.gov (ID NCT05479097).

## Funding

Research reported in this publication was supported by the Fogarty International Center of the National Institutes of Health under award number R21TW011891. During the research period, TV was funded in part by the NIH Medical Scientist Training Program grant T32 GM140935. The content is solely the responsibility of the authors and does not necessarily represent the official views of the National Institutes of Health. The NIH was not involved in study design or execution, nor the decision to submit for publication.

## Authorship contributions

SD assumed the lead role in study design, clinical algorithm development, data review and descriptive analysis, statistical analysis, manuscript drafting and editing, and contributed to literature review. AC led mHealth application design and contributed to study design and manuscript drafting and editing. IA contributed to statistical analysis, manuscript drafting, and editing. GO contributed to literature review, manuscript drafting and editing. AB and PP performed mHealth application testing and contributed to manuscript drafting and editing. EW performed mHealth application testing and contributed to manuscript drafting and editing. VA performed mHealth application testing and CHW training and contributed to manuscript drafting and editing. JA led literature review and contributed to study design, mHealth application testing, CHW training, and manuscript drafting and editing. DD contributed to study design, performed mHealth application testing, and contributed to manuscript drafting and editing. CP provided feedback during mHealth application development and contributed to manuscript drafting and editing. YL and RT contributed to study design and clinical algorithm development, provided feedback during mHealth application development, were involved in data acquisition as physician consultants, and contributed to manuscript drafting and editing. TV led study coordination, contributed to study design, collected and analyzed qualitative data, and contributed to manuscript drafting and editing.

## Disclosure of interest

In addition to the NIH funding support noted above, SD reports honoraria and support from the American Academy of Family Physicians (AAFP) for attendance and travel to the AAFP Global Health Summit in 2023 and 2025 to deliver presentations related to this project. TV reports funding from several US federal sources and universities to support her PhD studies during the time this project was being carried out, as well as scholarship support from the AAFP to attend the AAFP Global Health Summit in 2025 to present on this project along with SD.

## Data Availability

The datasets generated and/or analyzed during the current study are not publicly available as such sharing outside of the research team was not part of the research protocols approved by ethical oversight committees for this project, but may be made available upon reasonable request from the corresponding author with IRB permission.

## Notes

### Clinical Trial

NCT05479097

### Author Declarations

This research was carried out in accordance with the Declaration of Helsinki. The study was approved by the University of Wisconsin Institutional Review Board (ID # 2022-0794, initial approval 07/02/2022) and by the San Lucas Healthcare Committee (initial approval 09/19/2020), an interdisciplinary, international group which provides local ethics oversight to healthcare projects. Eligible patients provided written informed consent prior to participating in the study. Study procedures, potential risks and benefits were explained to potential participants by the community health worker (CHW) in Spanish and/or Kaqchikel depending on the patient's preferred language, and any question answered. A copy of the study information and consent form in Spanish (as patients are generally only literate in Spanish even if they primarily speak Kaqchikel) was provided to the participant. If the CHW obtaining consent was unsure about the patient's ability to provide informed consent, one of the study supervising physicians was consulted to assess the participant and determine if they could provide consent. This study was registered with ClinicalTrials.gov (ID NCT05479097).

